# Bicentric evaluation of stabilizing sampling tubes for assessment of monocyte HLA-DR expression in clinical samples

**DOI:** 10.1101/2021.05.12.21256300

**Authors:** Sarah Hamada, Robin Jeannet, Morgane Gossez, Martin Cour, Laurent Argaud, Bruno Francois, Thomas Daix, Fabienne Venet, Guillaume Monneret

**Affiliations:** Hospices Civils de Lyon, Edouard Herriot Hospital, Immunology Laboratory, 69437 Lyon, France; Inserm CIC 1435 Dupuytren Teaching Hospital, 87000 Limoges, France; UMR CNRS 7276, INSERM 1262, Faculty of Medicine, University of Limoges, 87000 Limoges, France; Centre International de Recherche en Infectiologie (CIRI), Inserm U1111, CNRS, UMR5308, Ecole Normale Supérieure de Lyon, Université Claude Bernard-Lyon 1, Team ‘NLRP3 inflammation and immune response to sepsis’, Lyon, France; Hospices Civils de Lyon, Edouard Herriot Hospital, Medical intensive Care Department, 69437 Lyon, France; Dupuytren Teaching Hospital, Medical-Surgical Intensive Care Unit, 87000 Limoges, France; EA 7426 “Pathophysiology of Injury-Induced Immunosuppression” (Université Claude Bernard Lyon 1 - Hospices Civils de Lyon - bioMérieux), Joint Research Unit HCL-bioMérieux, 5 place d’Arsonval, 69003, Lyon, France

## Abstract

**Background:** Diminished expression of human leukocyte antigen DR on circulating monocytes (mHLA-DR), measured by standardized flow cytometry procedure, is a reliable indicator of immunosuppression in severely injured intensive care unit patients. As such, it is used as stratification criteria in clinical trials evaluating novel immunostimulating therapies. Pre-analytical constraints relative to the short delay between blood sampling and flow cytometry staining have nevertheless limited its use in multicentric studies. The objective of the present work was to compare mHLA-DR expression between whole blood samples simultaneously drawn in EDTA or Cyto-Chex^®^ BCT tubes.

**Methods:** In 2 university hospitals, mHLA-DR was assessed in fresh whole blood from septic patients (n = 12) and healthy donors (n = 6) simultaneously sampled on EDTA and Cyto-Chex^®^ BCT tubes. Staining was performed immediately after sampling and after blood storage at room temperature.

**Results:** We observed the remarkable stability of mHLA-DR results when blood was collected in Cyto-Chex^®^ BCT tubes (until 48-72 h). On baseline values, despite good correlation between tubes (r = 0.98, p< 0.001), mHLA-DR expression was systematically lower with Cyto-Chex^®^ BCT.

**Conclusion:** The present reports confirms the great potential of Cyto-Chex^®^ BCT tubes to delay mHLA-DR staining in centers without rapid access to flow cytometry facilities. However, a 30 % gap exists between results obtained with EDTA and Cyto-Chex^®^ BCT tubes. As current thresholds for clinical decisions were obtained with EDTA samples, further studies are needed to confirm clinical thresholds with Cyto-Chex^®^ BCT tubes.

## Introduction

Sepsis, including viral infections such as COVID-19, is characterized by a complex immune response that varies over time, with the concomitant occurrence of both pro- and anti-inflammatory mechanisms (1). As a resultant, some septic patients enter a stage of protracted immunosuppression (2). This paved the way for tailored immunostimulation for patients presenting with the most altered or persistent immune functions (3). To date, the diminished expression of the human leucocyte antigen-DR on circulating monocytes (mHLA-DR) is consensually accepted as a reliable marker of immunosuppression in sepsis and severe injuries (4,5). It has been used to guide therapy in randomized clinical trials or complicated clinical cases (6-9). The measurement of mHLA-DR is based on a standardized flow cytometry protocol (10). This technique, published in 2005 by a group of experts, is based on the use of calibrated beads (i.e. BD Quantibrite anti-HLA-DR) with known and qualibrated amounts of fluorochrome to convert means of fluorescence intensities to numbers of antibodies bound per cell (AB/C). As this simple protocol requires only 2 monoclonal antibodies (anti-HLA-DR and anti-CD14 Abs), it is accessible to most inclusion centers with flow facilities. Inter-laboratory assessment between centers equipped with cytometers from different manufacturers provided excellent result (11). However strict pre-analytical conditions are mandatory such as staining within 1.5 h after blood collection or within 4h if storage at + 4°C in order to avoid non-specific rise of mHLA-DR expression over time (12). These constraints have limited a larger use of mHLA-DR in clinical practice. To circumvent these pre-analytical hurdles, blood collection could be performed directly in stabilizing tubes such as Cyto-Chex^®^ BCT tubes (rather than EDTA ones). These are direct blood collection tubes for the collection and storage of whole blood specimens for immunophenotyping of white blood cells by flow cytometry. They supposedly maintain cellular morphology and surface antigen expression, up to 72h post-collection even if stored at room temperature.

Recently, low mHLA-DR has been proposed as enrollment criteria in sepsis multicentric clinical trials (e.g., anti-PD-L1, #NCT 02576457). In this intended use, a strict reproducibility of values is essential to meet regulatory requirements. To this end, an extensive characterization of assay performance has been conducted according to the New York State Department of Health (NYSDOH) requirements (5). Results, obtained with standardized staining protocol (cited above) and Cyto-Chex^®^ BCT tubes demonstrated (i) the stability of mHLA-DR values at room temperature (until 72 h after sampling) and (ii) the adequacy of this approach (e.g., precision, accuracy, lab-to-lab comparison) to fulfill requirements for regulatory approval (5). Nevertheless, the strict comparison of mHLA-DR values between blood sampled in EDTA versus Cyto-Chex^®^ BCT tubes was not performed in this validation study. As mHLA-DR clinical thresholds have all been determined in blood collected in EDTA tubes, validation of the comparability of the results obtained between these 2 types of tubes remained to be performed.

In order to complete the work of Quadrini and colleagues (5), the objective of the present work was to compare mHLA-DR values from blood simultaneously collected in EDTA and Cyto-Chex^®^ BCT tubes and to confirm over time stability of mHLA-DR expression with Cyto-Chex^®^ BCT tubes.

## Methods

### Whole blood samples

Samples of whole blood were simultaneously collected both in EDTA anticoagulant and Cyto-Chex^®^ BCT tubes (Streck, La Vista, NE) from 12 COVID-19 patients: n = 6, from medical intensive Care Department, Hôpital E. Herriot, Lyon, F (RICO study, (IRB #: IORG0009918, agreement number 2020-A01079-30, ClinicalTrials number: NCT04392401) and n = 6, from medical intensive Care Department, Hôpital Dupuytren, Limoges, F (ILIAD-7 study, ClinicalTrials number: NCT04379076)) and from 6 healthy volunteers (HV) provided by the “Etablissement Français du Sang (EFS)”. Blood donors’ personal data were totally anonymized before transfer to our research laboratory.

### Cell Preparation and Flow Cytometric Analysis

mHLA-DR determination was performed on both types of tube using standardized protocol as already described (5,10,11). Briefly, whole blood (25 µL) was stained with 10 µL of QuantiBrite HLA-DR/Monocyte mixture (QuantiBrite anti-HLA-DR PE (clone L243)/Anti-monocytes (CD14) PerCP-Cy5.5 (clone MUP9), Becton Dickinson San Jose, CA, USA) at room temperature for 30 min in a dark chamber. Samples were then lysed using the FACS Lysing solution (Becton Dickinson) for 15 min. After a washing step, cells were analysed on cytometer Navios (BeckmanCoulter). Monocytes were first gated out from other cells on the basis of CD14 expression and mHLA-DR expression was then measured on their surface (mono-parametric histogram) as median of fluorescence intensity related to the entire monocyte population (as recommended by manufacturer). These results were then transformed in AB/C (number of antibodies bound per cell) thanks to calibrated PE-beads (BD QuantiBriteTM-PE Beads, Becton Dickinson) that were run on each flow cytometer.

mHLA-DR staining (both EDTA and tubes Cyto-Chex^®^ BCT) was performed immediately after sampling and repeated on blood stored at room temperature after 6 hours, 24 hours, 48-72 hours on both EDTA and Cyto-Chex BCT tube sand after 168 hours (Limoges only with Cyto-Chex^®^ BCT tubes)

### Statistical analysis

Results are presented as means +/- standard deviations. Correlations were evaluated using the Spearman correlation coefficient and Bland Altman representation. Non parametric Wilcoxon paired-test was used to compare values from EDTA and Cyto-Chex^®^ BCT tubes. The statistical analyses were performed using GraphPad Prism 8.0.1. A p value below 0.05 was considered as significant.

## Results

### Over time stability

Results are presented in Figure 1. As previously shown, we observed a massive elevation of mHLA-DR expression in EDTA tubes stored at room temperature (Figure 1A). The phenomenon was similarly observed in healthy donors and COVID-19 patients with same magnitude as we observed more than 100 % enhancement above baseline when mHLA-DR was measured after 48-72 h of blood storage at RT (Figure 1B). In contrast, results obtained with Cyto-Chex^®^ BCT tubes were remarkably stable both in heathy donors and patients. Indeed, most results elevation after blood storage remained below 20 % from baseline (Figure 1B) which was in agreement with acceptance criteria by NYSDOH. Individual values depicted same stability (Figure 1C).

**Figure 1.**
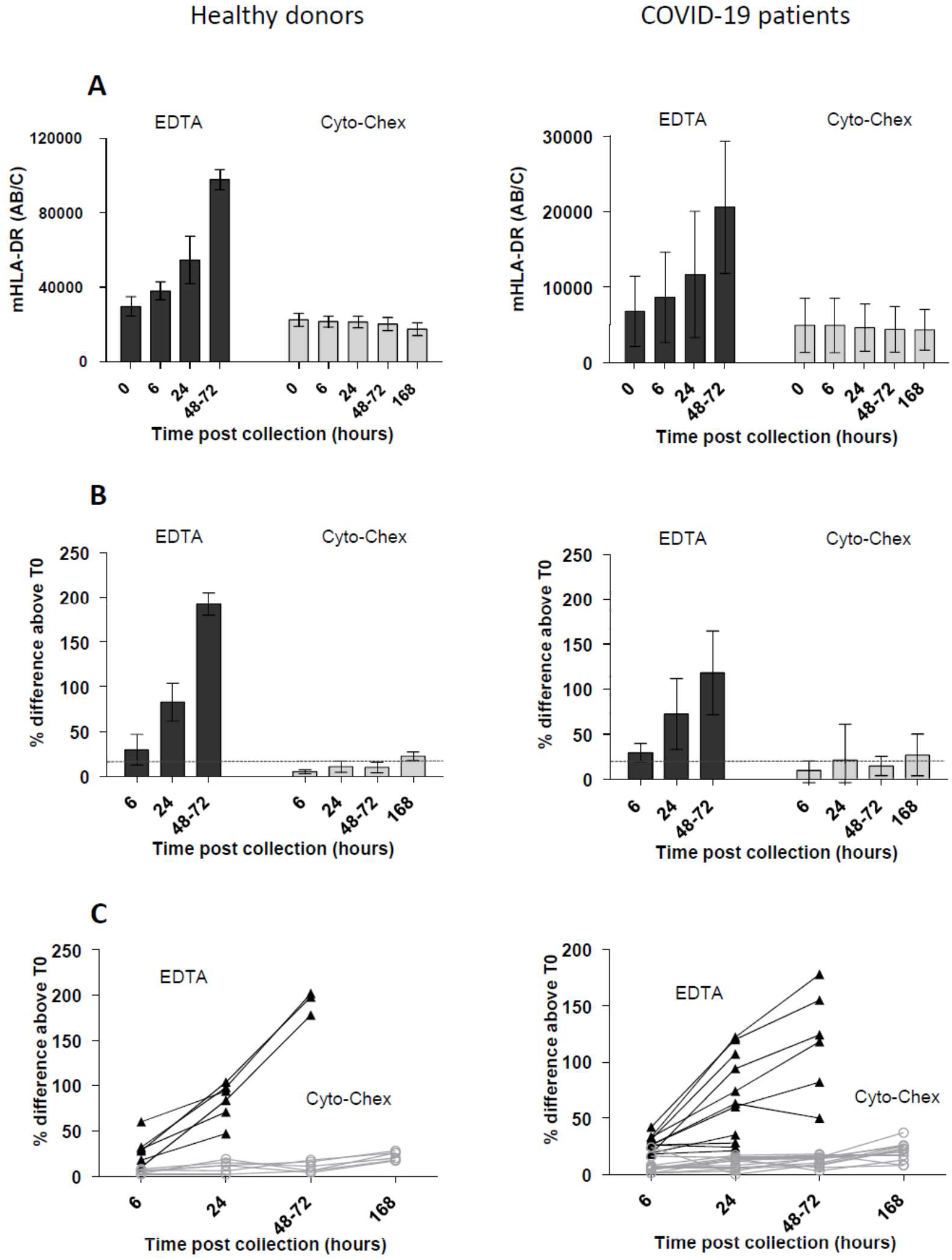
Overtime stability at room temperature of monocyte HLA-DR expression. Comparison of values in septic patients (n = 12) and healthy donors (n = 6) simultaneously sampled on EDTA (black bars and triangles) and Cyto-Chex^®^ BCT tubes (grey bars and open circles). Results are expressed as **(A)** numbers of antibodies bound per cell (AB/C, means +/- SD), **(B)** % difference above t = 0 (i.e., immediate staining after blood collection, means +/- SD), **(C)** individual data (one patient with very low mHLA-DR value (< 500 AB/C) is not represented).

### EDTA vs Cyto-Chex^®^ BCT values immediately after sampling

As mHLA-DR clinical thresholds have been determined on monocytes collected in EDTA tubes, we next investigated whether both blood sampling tubes may provide comparable mHLA-DR results. For this purpose, we only considered mHLA-DR values obtained upon sampling (i.e., without blood storage). While values were nicely correlated (r = 0.98, p< 0.001, Figure 2A); Bland Altman plot indicated a systematic bias between protocols (Figure 2B). On average, Cyto-Chex^®^ BCT tubes provided mHLA-DR values approximately 30 % lower than those obtained with EDTA (Figure 2B). This difference was significant and systematically observed (p < 0.0005, non-parametric wilcoxon paired test, Figure 2C).

**Figure 2.**
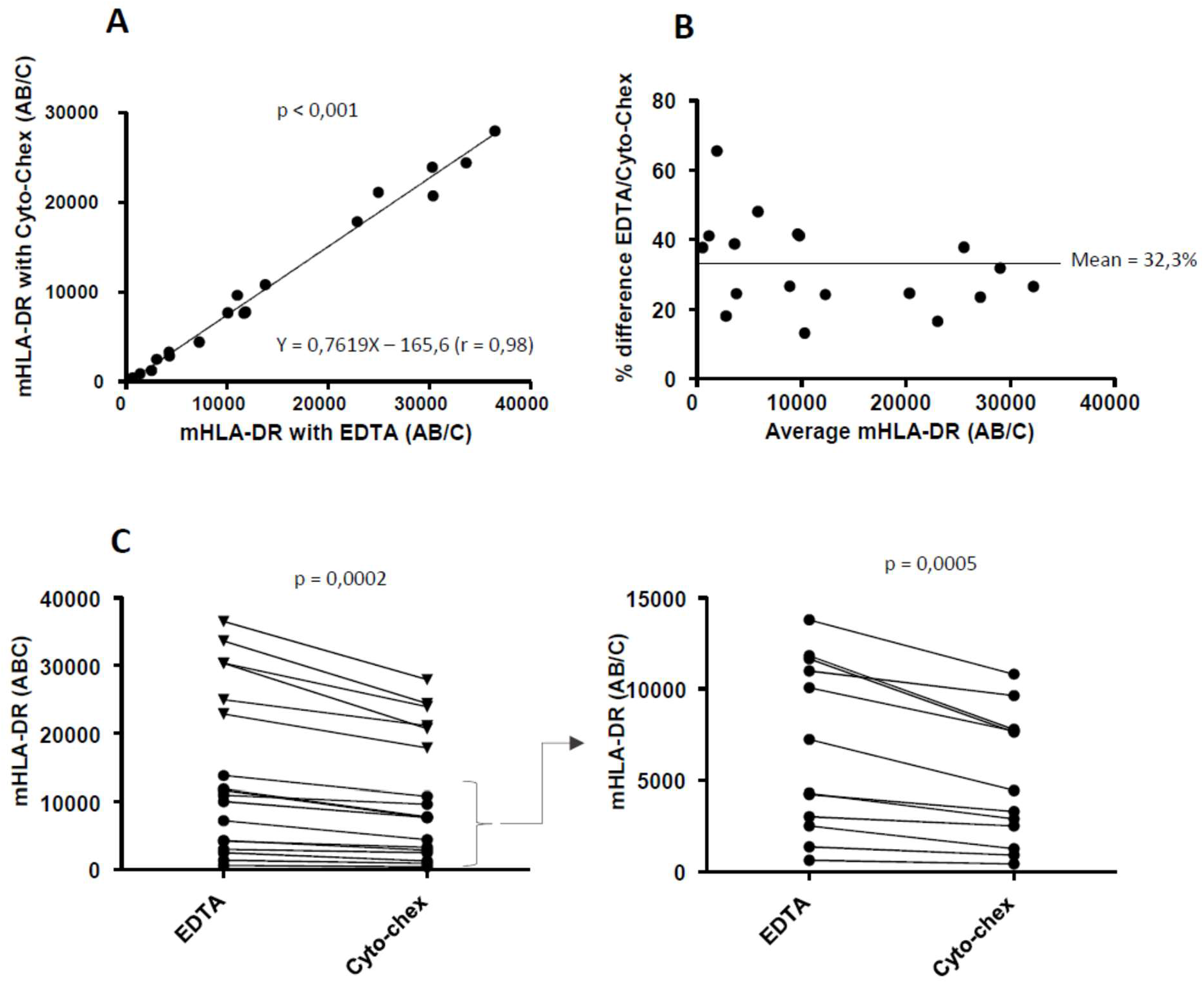
Comparison of monocyte HLA-DR results at baseline between EDTA and Cyto-Chex^®^ BCT tubes. **(A)** Correlation, Spearman correlation coefficient is given. **(B)** Bland-Altman plots, dashed line represents mean of differences between tubes **(C)** individual paired samples (left: all samples, triangles: healthy donors, circles: COVID-19 patients, right: focus on values < 15 000 AB/C, statistics: Wilcoxon paired test). Values from septic patients (n = 12) and healthy donors (n = 6) simultaneously sampled on EDTA and Cyto-Chex^®^ BCT tubes and immediately stained for mHLA-DR were analyzed.

## Discussion

The World Health Organization (WHO) recently recognized sepsis as a global health priority by adopting a resolution to improve the prevention, diagnosis and management of this deadly disease (13). Sepsis is defined as a life-threatening organ dysfunction due to a dysregulated response to infections (14) and remains the leading cause of death in intensive care unit (15). After a short pro-inflammatory phase, some septic patients enter a stage of profound immunosuppression (2). This contributes to the body’s incapacity to clear the initial infectious focus (16). This is also illustrated in those severely injured patients by reactivation of dormant viruses (CMV or HSV) or infections due to pathogens, including fungi, which are normally pathogenic solely in immunocompromised hosts (17). As nearly all immune functions are deeply compromised, defects in immune surveillance contribute to worsening outcome in patients who survived initial resuscitation. Consequently, new promising therapeutic avenues are currently emerging such as adjunctive immunostimulation (IFN-g, IL-7, anti-PD1/L1 antibodies) for the most immunosuppressed patients (6,18,19). Nevertheless, as there is no clinical sign of immune dysfunctions, the prerequisite for such therapeutic intervention relies on our capacity to perform immunophenotyping in order to identify the subgroup of patients who could benefit from immunostimulation.

To date, decreased mHLA-DR expression appears as the most robust biomarker in the field (20). As such, low mHLA-DR associates with increased nosocomial infection occurrence and mortality (4). Monocytes expressing low mHLA-DR are indeed inefficient to present antigen to T lymphocytes, to produce inflammatory cytokines and to mount efficient immune response against secondary infections or to clear initial infection causing sepsis. Most importantly, mHLA-DR has been selected as an enrollment criteria for randomized clinical trial aimed at assessing various immunostimulant molecules in sepsis (7). However, in case of multicentric trials, the absolute precondition is the absence of inter-laboratory variability between results obtained from different labs. As explained above, strict pre-analytical conditions related to nonspecific rise of mHLA-DR in EDTA tubes upon blood storage prevented its wide use in RCT due to the necessity to have rapid access to flow cytometry facilities in all centers. This may have a major impact on potential clinical interpretation. Indeed, in the present study, we confirmed that most patients’ values were initially below 8 000 AB/C (i.e., threshold indicative of immunosuppression) but were found above > 10 000 AB/ C after 24 h of EDTA blood storage, which would indicate acceptable level of immune competence in intensive care unit patients (10). In this context, the provision of Cyto-Chex^®^ BCT supposed to stabilize cell surface marker expression appeared as a potential major step for multicenter deployment of mHLA-DR staining. Using these tubes and Quantibrite staining kit, Quadrini and colleagues (5) recently made a crucial contribution to the field by demonstrating (i) the stability of mHLA-DR expression up to 72 h after blood collection and (ii) the adequacy of the approach to fulfill criteria for NYSDOH approval. However, the question of comparison of mHLA-DR values obtained with EDTA vs Cyto-Chex^®^ BCT tubes, although being of major importance when designing clinical thresholds, remained unresolved.

In the present study and as described by Quadrini et al (5), we confirmed the remarkable stability of mHLA-DR expression up to 72 h after blood collection in Cyto-Chex^®^ BCT tubes while mHLA-DR expression significantly increased in EDTA tubes. Most importantly, we showed that Cyto-Chex^®^ BCT values were systematically 30 % below those obtained with EDTA samples. We hypothesized that reagents used in Cyto-Chex^®^ BCT to stabilize surface protein expression may partly hide some HLA-DR epitopes. This systematic gap between mHLA-DR values may be of crucial importance. For example, in clinical trial evaluating GM-CSF in sepsis, Meisel et al (7) used a clinical threshold at 8000 AB/C to include patients. Based on this threshold, 3 patients from the present study who presented with mHLA-DR values > 10 000 AB/C when measured in EDTA tubes (i.e. close to normal mHLA-DR values (10) would have been enrolled based on mHLA-DR measured in Cyto-Chex^®^ BCT tubes (i.e., 7784, 7634, 7659 AB/ C with Cyto-Chex^®^ BCT instead of 11816, 11645, 10066 respectively with EDTA). Taking into account the good correlation between tubes but to correct for this difference, we could estimate that, this threshold would approximately have to be lowered to 5 600 AB/C with blood samples drawn in Cyto-Chex^®^ BCT tubes (i.e., 30 % of 8 000). Nevertheless, the precise clinically pertinent inclusion threshold of mHLA-DR obtained with Cyto-Chex^®^ BCT tubes would have to be confirmed in a prospective clinical study.

To sum up, we confirmed the great potential of Cyto-Chex^®^ BCT tubes for mHLA-DR measurements to delay cells staining in centers without rapid access to flow cytometry facilities. The use of such tubes could represent a quantum leap in multicentric RCT involving mHLA-DR assessment. However, a 30 % gap exists, between values obtained in Cyto-Chex^®^ BCT versus EDTA tubes. As clinical thresholds of mHLA-DR to define ICU patients’ immune status have been obtained with EDTA-sampled blood, this difference should be kept in mind when designing next clinical trials in order to adapt clinical threshold for decision making. A study including a large number of septic patients to simultaneously compare Cyto-Chex^®^ BCT and EDTA tubes would be of major interest to establish concordant clinical thresholds between both protocols.

## Data Availability

Data available upon request

## Acknowledgments

This work was supported by Hospices Civils de Lyon and Lyon 1 University (Lyon, F); and Dupuytren Teaching Hospital (Limoges, F).

